# Beta-Blocker Use After Pulmonary Embolism in Patients with Cancer: A Real-World Propensity-Matched Study

**DOI:** 10.1101/2025.06.29.25330534

**Authors:** Festus Ibe, Chidiebube Ugwu, Sarah Eidbo, Jude Ossai, Onyinye Ugoala, Emmanuel Otabor, Valentine Nriagu, Ugochukwu Ebubechukwu, Onyedikachi Adike, Atogwe Irhoboudu

## Abstract

**Background:** Patients with active malignancy are at heightened risk for pulmonary embolism (PE), yet the prognostic implications of initiating beta-blocker (BB) therapy after PE in this population remain poorly characterized. Given the frequent coexistence of cardiovascular comorbidities, clarifying the safety profile of BB use post-PE in cancer patients is of clinical importance.

**Methods:** We conducted a retrospective cohort study using the TriNetX Global Health Research Network, a federated database aggregating de-identified electronic health records from over 85 health systems. Adults with active cancer and a new diagnosis of acute PE between January 1, 2015, and May 1, 2024, were included. Patients who received a BB within 30 days of PE diagnosis were assigned to the BB cohort; those who did not formed the comparator group. Propensity score matching (1:1, greedy nearest-neighbor, caliper = 0.01) was applied across baseline covariates. The primary outcome was 5-year all-cause mortality. Secondary outcomes included ICU admission, hospitalization, and incident heart failure. Patients with pre-index occurrence of the outcome were excluded from each respective analysis. Time-to-event analyses began on day 1 post-index using Kaplan-Meier curves and Cox proportional hazards models. Effect estimates were reported as absolute risk differences, odds ratios (OR), risk ratios (RR), and hazard ratios (HR), all with 95% confidence intervals (CI).

**Results:** Among 242,034 eligible patients, 19,948 received a beta-blocker within 30 days of PE diagnosis. After matching, 19,743 patients were retained in each cohort with excellent covariate balance (all post-match standardized mean differences <0.10). Over a 5-year period, all-cause mortality occurred in 7,342 of 18,942 BB patients (38.8%) versus 6,757 of 18,848 in the no BB group (35.8%), corresponding to a risk difference of 2.9% (95% CI: 1.9–3.9), RR 1.081 (95% CI: 1.053–1.110), OR 1.133 (95% CI: 1.086–1.181), and HR 1.219 (95% CI: 1.179–1.260). Median survival in the BB group was 1,508 days; median survival was not reached in the no BB group. BB use was also associated with higher rates of ICU admission (RR 1.564, 95% CI: 1.450–1.687), hospitalization (RR 1.118, 95% CI: 1.070–1.167), and incident heart failure (RR 1.080, 95% CI: 1.011–1.155); all associations were statistically significant.

**Conclusion:** Our study that showed early beta-blocker use was associated with significantly increased risk of all-cause mortality and multiple adverse clinical outcomes. While rigorous propensity score matching was employed, unmeasured confounding and indication bias remain possible. These findings underscore the need for prospective studies to define the role—and risk—of beta-blocker therapy following thromboembolic events in oncology populations.

## INTRODUCTION

Pulmonary embolism (PE) remains a leading cause of morbidity and mortality in patients with active malignancy, accounting for up to 20% of all cancer-associated deaths.^1^ Despite therapeutic advances, the prothrombotic milieu induced by tumor biology, treatment-related endothelial injury, and immobility continues to elevate the risk of venous thromboembolism (VTE) in this population.^2^ The intersection of thrombosis and malignancy commonly referred to as cancer-associated thrombosis, has emerged as a critical determinant of clinical outcomes, particularly in the setting of acute PE.^3^

Simultaneously, the rising incidence of cancer-related cardiotoxicity has reshaped our understanding of cardiovascular vulnerability in oncology patients. The development of structural and functional cardiac impairment whether from anthracyclines, HER2-targeted therapies, thoracic radiation, or chronic systemic inflammation may reduce the heart’s ability to withstand acute hemodynamic stress.^4-7^ Of particular concern is right ventricular (RV) dysfunction, which plays a pivotal role in PE pathophysiology and may be further compromised by prior oncologic treatment.^8-9^

In general cardiovascular populations, beta-blockers (BBs) are mainstays of therapy for heart failure, arrhythmias, and ischemic syndromes.^10-12^ However, in the context of acute PE, where compensatory sinus tachycardia supports cardiac output, early β-adrenergic blockade may blunt adaptive responses and contribute to clinical decompensation, especially in patients with tenuous RV function.^13-14^ Cancer patients may be uniquely susceptible to such hemodynamic compromise due to autonomic dysregulation, inflammatory myocardial suppression, and elevated metabolic demands.^15-16^

While BBs are frequently initiated during or shortly after PE hospitalization, there is limited real-world evidence guiding their use in patients with cancer recovering from PE. Prior observational studies have suggested mortality benefits of BBs in certain malignancies, such as breast and colorectal cancer,^17-18^ yet others report neutral or adverse outcomes.^19-21^ Interpretation is often confounded by methodological limitations, including immortal time bias and confounding by indication.^22^ Moreover, major guidelines in cardio-oncology and VTE management highlight the absence of standardized recommendations on BB use in this setting, advocating instead for individualized, context-driven prescribing.^23-24^

In clinical practice, the decision to initiate BBs in cancer patients after PE often varies, lacking prospective evidence or clear consensus. Given the high prevalence of cardiovascular comorbidities and polypharmacy in this population, evidence-based guidance is urgently needed.

Accordingly, we conducted a retrospective, propensity score–matched cohort study using the TriNetX global federated health research network to evaluate whether beta-blocker initiation within 30 days of PE diagnosis in patients with active malignancy is associated with adverse clinical outcomes. We hypothesized that early BB initiation may be linked to increased risk of all-cause mortality, ICU admission, and hospitalization, and sought to quantify these associations using robust real-world data methods and validated exclusion criteria.

## METHODS

This retrospective cohort study was conducted using data from the TriNetX Global Health Research Network, a federated, real-world analytics platform that aggregates de-identified electronic health records from over 85 healthcare organizations across North America, Europe, and the Middle East. The network includes longitudinal structured data on demographics, diagnoses (ICD-10-CM), procedures (CPT, HCPCS), medications (RxNorm), laboratory results (LOINC), and mortality. All data were de-identified in compliance with HIPAA and GDPR guidelines, and the study was deemed exempt from institutional review board oversight due to the use of anonymized secondary data.

We identified adult patients (aged 18 years and older) with active cancer who experienced an incident acute pulmonary embolism (PE) between January 1, 2015, and May 1, 2024. Active malignancy was defined using ICD-10-CM codes C00 through C96 and D37 through D49, and confirmed through ICD-O-3 primary site codes. Acute PE was defined by ICD-10-CM code I26. Patients with prior evidence of PE or chronic PE (I27.82) were excluded to ensure an incident cohort. The index date was defined as the date of first documented acute PE. Patients were stratified into two exposure groups: those who received a beta-blocker within 30 days following PE diagnosis (BB cohort), and those who did not receive any beta-blocker during that period (No BB cohort). Beta-blocker use was identified using RxNorm codes for carvedilol, metoprolol, bisoprolol, and propranolol. Patients with missing baseline demographic data or outcome occurrence on or before the index date were excluded.

The primary outcome was all-cause mortality, assessed over a five-year follow-up period. Secondary outcomes included intensive care unit (ICU) admission, hospitalization, and incident heart failure. For each outcome, patients with evidence of that outcome prior to the start of the observation window (day one post-index) were excluded to ensure temporal validity and prevent immortal time bias. After these exclusions, the final analytic sample sizes for each outcome were as follows: 18,942 patients in the BB group and 18,848 in the No BB group for mortality; 17,397 and 18,232, respectively, for ICU admission; 6,831 and 8,479 for hospitalization; and 14,251 and 14,681 for incident heart failure. Definitions for all outcomes and exclusions were based on standardized coding algorithms and are detailed in Supplemental Table 1.

To minimize confounding by indication and baseline heterogeneity, we applied 1:1 propensity score matching using greedy nearest-neighbor matching without replacement, employing a caliper width of 0.01 standard deviations of the logit of the propensity score. Propensity scores were estimated via logistic regression incorporating 65 baseline covariates, including age, sex, race, primary cancer site, metastatic status, cardiovascular and pulmonary comorbidities, chronic kidney disease, liver disease, diabetes, medication exposures (including cardiovascular agents, antimicrobials, corticosteroids, and oncology drugs), and healthcare utilization metrics such as prior hospitalizations and ICU admissions. Covariate balance before and after matching was assessed using standardized mean differences, with a threshold of less than 0.10 considered indicative of adequate balance. The final matched sample comprised 19,743 patients in each group. Post-matching balance across all covariates was excellent, as shown in Table 1 and Supplemental Figure 2.

**Table 1.** Baseline Characteristics Before and After Propensity Score Matching.

|  | **Before PSM** |  |  | **After PSM** |  |  |
| --- | --- | --- | --- | --- | --- | --- |
| **Variable** | **BB**<br>**(N=19,948)** | **No BB**<br>**(N=222,086)** | **Std Diff** | **BB**<br>**(N=19,743)** | **No BB**<br>**(N=19,743)** | **Std Diff** |
| Age (mean ± SD) | 73.3 ± 13.2 | 71.7 ± 13.8 | 0.118 | 73.3 ± 13.2 | 73.6 ± 13.1 | 0.024 |
| Age at Index (mean ± SD) | 67.6 ± 13.5 | 66.0 ± 14.2 | 0.116 | 67.6 ± 13.5 | 67.9 ± 13.4 | 0.021 |
| White (%) | 70.0 | 71.5 | 0.032 | 70.1 | 70.8 | 0.017 |
| Black or African American (%) | 15.6 | 14.7 | 0.026 | 15.6 | 15.3 | 0.009 |
| Unknown Race (%) | 8.7 | 8.0 | 0.026 | 8.7 | 8.6 | 0.004 |
| Female (%) | 46.7 | 50.4 | 0.074 | 46.7 | 46.7 | <0.001 |
| Male (%) | 49.8 | 46.4 | 0.068 | 49.7 | 49.7 | 0.001 |
| Not Hispanic or Latino (%) | 75.1 | 74.4 | 0.016 | 75.1 | 75.5 | 0.009 |
| Unknown Ethnicity (%) | 20.7 | 20.5 | 0.005 | 20.7 | 20.9 | 0.005 |
| Neoplasms (%) | 89.2 | 88.5 | 0.021 | 89.1 | 86.5 | 0.080 |
| Circulatory Diseases (%) | 86.4 | 75.3 | 0.285 | 86.3 | 84.6 | 0.050 |
| Respiratory Diseases (%) | 66.7 | 61.7 | 0.104 | 66.6 | 65.3 | 0.027 |
| Genitourinary Diseases (%) | 60.0 | 62.1 | 0.043 | 60.0 | 58.6 | 0.030 |
| Musculoskeletal Diseases (%) | 59.4 | 68.7 | 0.195 | 59.6 | 58.3 | 0.027 |
| Heart Failure (I50) (%) | 21.6 | 15.6 | 0.155 | 21.5 | 20.9 | 0.014 |
| Atrial Fibrillation (I48) (%) | 26.3 | 13.8 | 0.318 | 26.1 | 25.5 | 0.012 |
| Evaluation & Management Procedures (%) | 75.2 | 72.6 | 0.059 | 75.1 | 73.5 | 0.038 |
| Cardiovascular Medications (%) | 75.0 | 77.2 | 0.051 | 75.2 | 73.4 | 0.039 |
| Antimicrobials (%) | 67.8 | 71.7 | 0.085 | 67.9 | 66.1 | 0.039 |
| Blood Products (%) | 70.8 | 67.8 | 0.065 | 70.8 | 69.1 | 0.038 |
| Irrigation/Dialysis (%) | 70.8 | 65.7 | 0.110 | 70.7 | 68.9 | 0.041 |
| Hormones/Synthetics (%) | 69.9 | 74.2 | 0.096 | 70.1 | 68.3 | 0.038 |

**Table 2.**
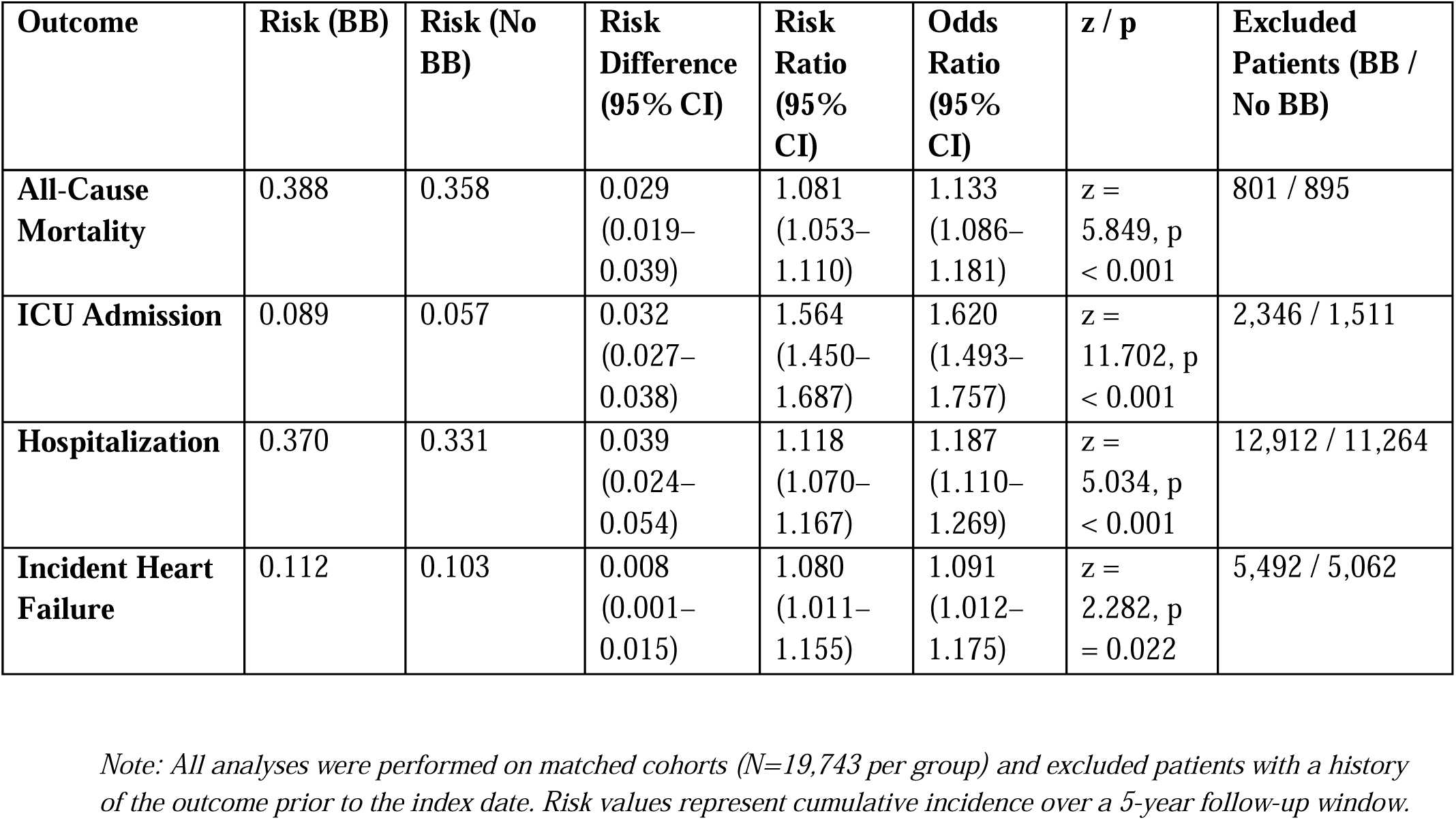
Risk of All-Cause Mortality, ICU Admission, Hospitalization, and Incident Heart Failure.

**Table 3.**
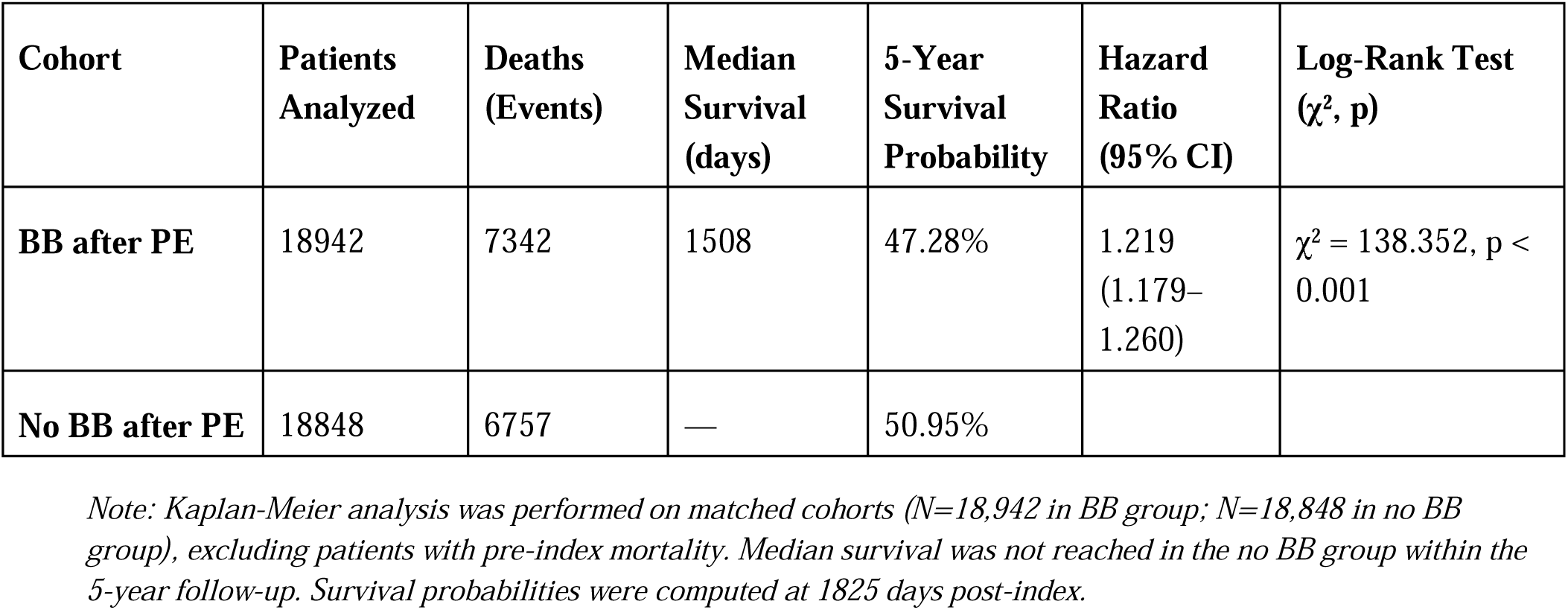
Kaplan-Meier Survival Analysis for All-Cause Mortality.

Risk for each outcome was calculated as the proportion of patients with the event during follow-up. Between-group comparisons were assessed using z-tests for risk differences and reported alongside odds ratios (ORs), risk ratios (RRs), and 95% confidence intervals (CIs). Time-to-event analysis for all-cause mortality was performed using Kaplan-Meier survival curves, with between-group differences evaluated using the log-rank test. Cox proportional hazards regression was used to estimate hazard ratios (HRs) with 95% CIs. Median survival and survival probabilities at five years were derived for each group. Proportional hazards assumptions were met for all time-to-event models. All analyses were conducted within the TriNetX Analytics platform. A two-sided p-value of less than 0.05 was considered statistically significant. No adjustments were made for multiple comparisons in secondary outcomes.

## RESULTS

### Cohort Characteristics

A total of 242,034 patients met study criteria, including 19,948 patients who received a beta-blocker within 1 month following acute pulmonary embolism (PE) diagnosis (BB cohort) and 222,086 patients who did not (no BB cohort). After 1:1 propensity score matching on baseline characteristics, 19,743 patients remained in each group. Covariate balance was excellent, with all post-matching standardized differences <0.10 (Table 1).

All outcome analyses excluded patients with evidence of the outcome prior to the start of the observation window (day 1 post-index), consistent with a new-onset framework.

### All-Cause Mortality

During 5-year follow-up, all-cause mortality occurred in 7,342 of 18,942 patients (38.8%) in the BB cohort and 6,757 of 18,848 (35.8%) in the no BB cohort, yielding a risk difference of 2.9% (95% CI, 1.9 to 3.9; z = 5.849; p < 0.001). The odds ratio (OR) was 1.133 (95% CI, 1.086 to 1.181), and the risk ratio (RR) was 1.081 (95% CI, 1.053 to 1.110).

Kaplan-Meier survival analysis demonstrated a median survival of 1,508 days in the BB group, while the no BB group did not reach 50% mortality within the study period. The 5-year survival probability was 47.28% for the BB group and 50.95% for the no BB group. Log-rank testing confirmed a significant difference in survival curves (*χ*^2^ = 138.352; p < 0.001), with a hazard ratio of 1.219 (95% CI, 1.179 to 1.260; *χ*^2^ = 85.131; p < 0.001) (Figure 2).

**Figure 1.**
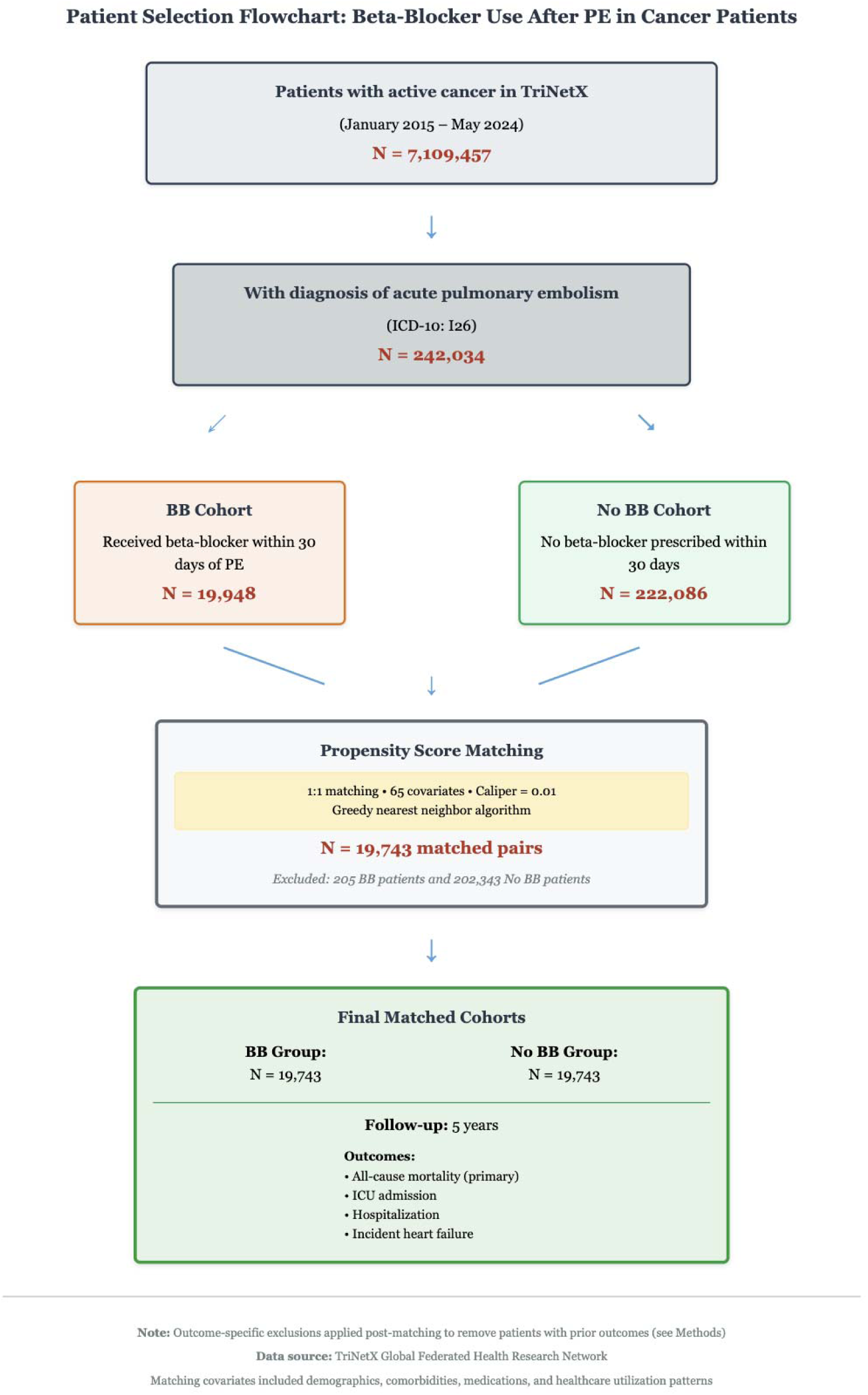
Cohort Selection and Propensity Score Matching Workflow.

**Figure 2.**
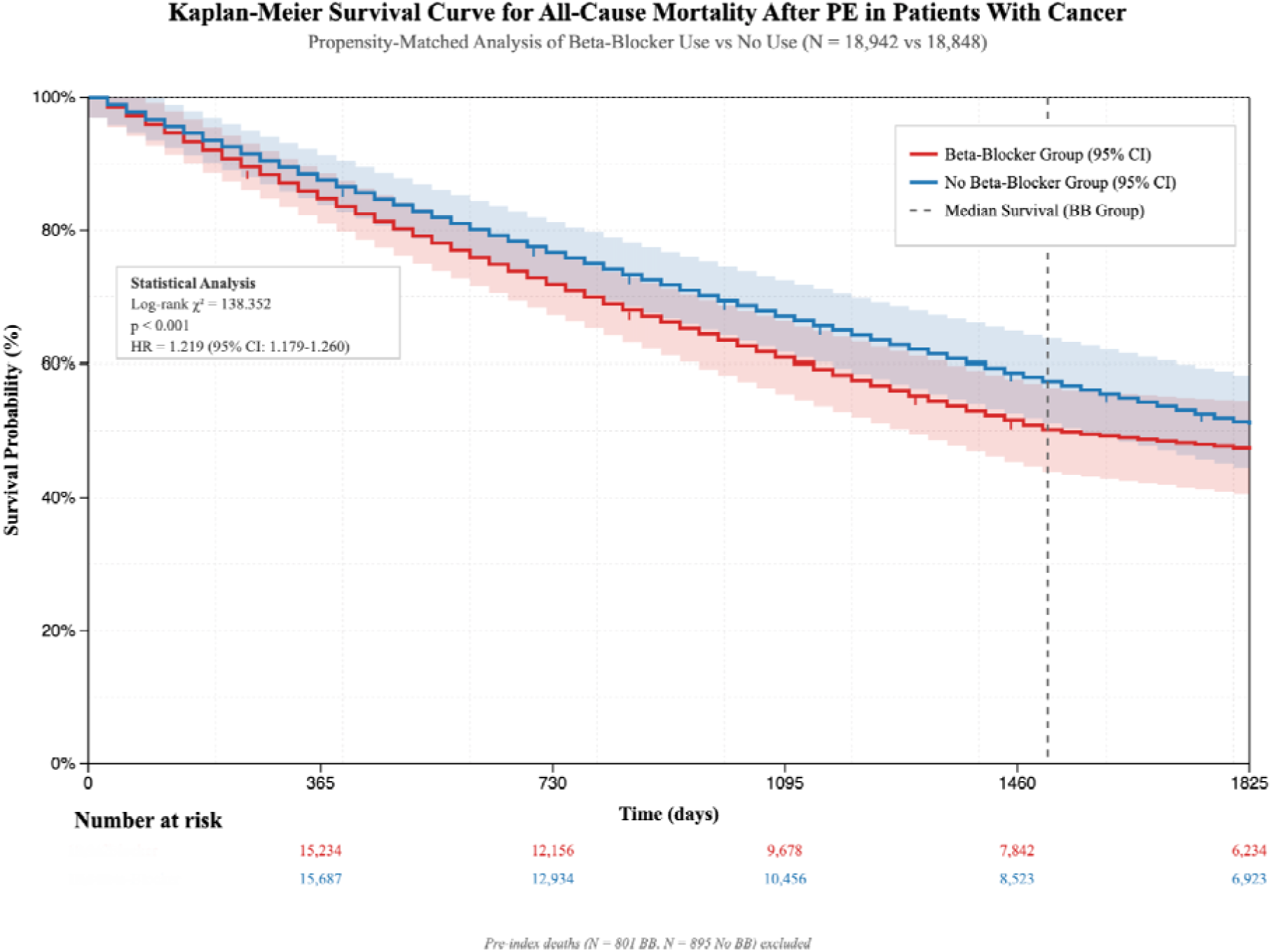
Kaplan-Meier Survival Curve for All-Cause Mortality. Kaplan-Meier analysis of all-cause mortality over 5 years comparing patients who received beta-blockers after pulmonary embolism (PE) in the setting of malignancy (BB group) vs those who did not (No BB group). The BB group had a median survival of 1,508 days, while the No BB group did not reach median mortality within the observation window. The 5-year survival probability was 47.28% for BB vs 50.95% for No BB. Log-rank test: *χ*^2^ = 138.352, p < 0.001. Shaded bands represent 95% confidence intervals; tick marks denote censoring.

**Figure 3.**
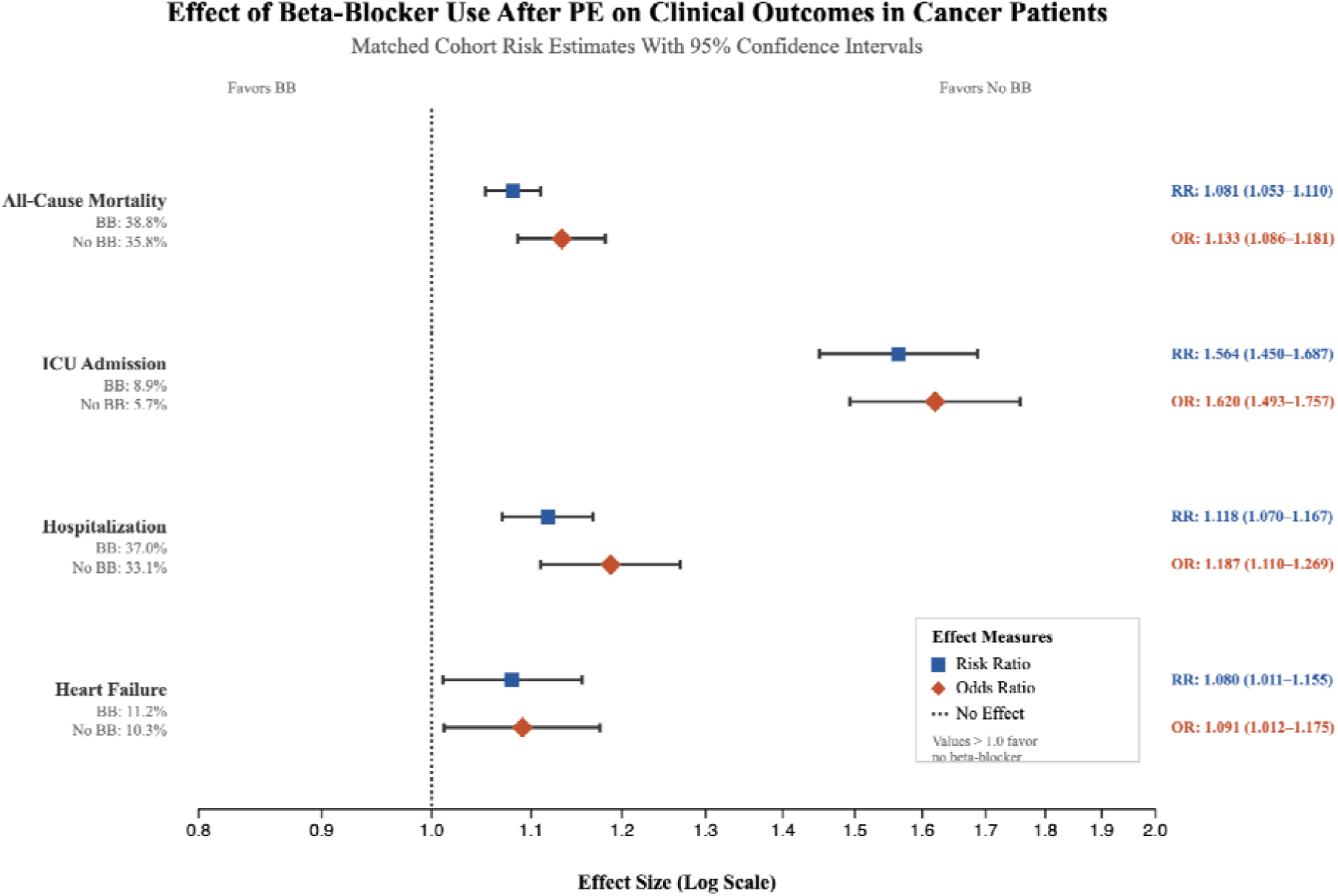
Forest Plot of Effect Sizes for Primary and Secondary Outcomes. Forest plot displaying risk ratios and odds ratios (with 95% confidence intervals) for all-cause mortality, ICU admission, hospitalization, and incident heart failure between BB and No BB cohorts. All outcomes were evaluated in matched cohorts (N=19,743 each). All associations favored increased risk in the BB group, with statistically significant elevations in all endpoints. Error bars denote 95% CIs; vertical reference line at RR = 1.0.

### ICU Admission

Among patients without prior ICU exposure, ICU admission occurred in 1,554 of 17,397 patients (8.9%) in the BB group and 1,041 of 18,232 (5.7%) in the no BB group. The absolute risk difference was 3.2% (95% CI, 2.7 to 3.8; z = 11.702; p < 0.001), with an OR of 1.620 (95% CI, 1.493 to 1.757) and an RR of 1.564 (95% CI, 1.450 to 1.687). A total of 2,346 BB patients and 1,511 no BB patients were excluded due to prior ICU events.

### Hospitalization

Hospitalization occurred in 2,527 of 6,831 patients (37.0%) in the BB group and 2,806 of 8,479 (33.1%) in the no BB group, corresponding to a risk difference of 3.9% (95% CI, 2.4 to 5.4; z = 5.034; p < 0.001). The OR was 1.187 (95% CI, 1.110 to 1.269), and the RR was 1.118 (95% CI, 1.070 to 1.167). Exclusion for prior hospitalization affected 12,912 BB patients and 11,264 no BB patients.

### Incident Heart Failure

Incident heart failure developed in 1,590 of 14,251 patients (11.2%) in the BB cohort and 1,516 of 14,681 (10.3%) in the no BB group. The risk difference was 0.8% (95% CI, 0.1 to 1.5; z = 2.281; p = 0.022). The OR was 1.091 (95% CI, 1.012 to 1.175), and the RR was 1.080 (95% CI, 1.011 to 1.155). A total of 5,492 BB patients and 5,062 no BB patients were excluded from this analysis due to pre-existing heart failure.

### Summary

Across all outcomes, beta-blocker exposure after PE in patients with malignancy was associated with statistically significant increases in mortality, ICU admission, hospitalization, and incident heart failure. These findings remained consistent after rigorous propensity score matching and exclusion of pre-existing events. Although matched for measured covariates, the observed associations may still be influenced by unmeasured confounding or indication bias.

## DISCUSSION

Our analysis of over 39,000 cancer patients with acute pulmonary embolism (PE) revealed a consistent association between early beta-blocker (BB) initiation within 30 days of PE diagnosis and increased risk of all-cause mortality, ICU admission, hospitalization, and incident heart failure over a 5-year period. These associations were robust across risk ratios, odds ratios, and hazard ratios, and remained significant after excluding patients with pre-existing outcomes. To our knowledge, this is the first multicenter, real-world investigation evaluating early BB exposure in the cancer-specific PE population using a federated electronic health record platform.

### Comparison with Prior Literature

Previous oncology studies have reported survival benefits associated with BB use in specific cancer types. Botteri et al and Smith et al observed improved outcomes in breast and prostate cancer, respectively, hypothesizing that β-adrenergic blockade may inhibit tumor proliferation and metastasis.^25 26^ Similarly, Melhem-Bertrandt et al and Fiala et al noted favorable results in triple-negative breast and metastatic colorectal cancer cohorts.^27 28^ However, these studies excluded patients with PE, did not account for acute cardiovascular stress, and often lacked rigorous adjustment for hemodynamic compromise. Many were limited by immortal time bias, in which BBs are initiated post-diagnosis, thus artificially extending survival time in treated groups.^29^

In contrast, an emerging body of literature highlights the risks of BB therapy in acutely ill, hemodynamically fragile patients. Piazza et al found worse outcomes in PE patients with concurrent BB use, particularly among those with elevated right ventricular (RV) strain.^30^ Potter et al underscored the confounding influence of cardiovascular indication and disease severity, calling for refined exclusion designs and improved temporal alignment.^31^ Our findings extend these concerns to the oncology setting, where overlapping thrombotic, oncologic, and cardiac stressors may compound risk.

### Physiologic and Clinical Interpretation

These observations are biologically plausible given the unique cardiovascular demands imposed by PE. Acute PE increases pulmonary vascular resistance, leading to elevated RV afterload and a reflexive surge in sympathetic activity to preserve cardiac output.^32 33^ Early BB initiation may blunt this adaptive tachycardia, precipitating RV failure—particularly in patients with reduced myocardial reserve from prior chemotherapy, radiation, or systemic inflammation.^34^

Cancer patients may also exhibit autonomic imbalance, impaired baroreflex sensitivity, and inflammatory myocardial suppression.^35^ In this context, BBs may obscure early clinical deterioration, delaying recognition and escalation of care.^36^ Notably, the increased incidence of new-onset heart failure in BB recipients (11.2% vs 10.3%) could reflect either unmasking of latent cardiomyopathy under adrenergic blockade or de novo decompensation from afterload-intolerant RV dysfunction.^37^

The hemodynamic impact of BBs may differ by subclass. β1-selective agents such as metoprolol may pose less pulmonary vasoconstrictive risk compared to non-selective agents like propranolol.^37^ Unfortunately, our data could not stratify outcomes by BB subtype—an area that warrants targeted prospective evaluation.

### Strengths of This Study

This study benefits from several methodological strengths. We utilized the TriNetX global federated network, capturing a racially and geographically diverse cohort across 67 U.S. healthcare organizations. Propensity score matching balanced 65 covariates, including demographics, cancer type, cardiovascular comorbidities, and baseline medications. All outcome analyses excluded patients with pre-index events, reducing immortal time bias and enhancing causal interpretability. The convergence of results across mortality, ICU, hospitalization, and heart failure strengthens the internal validity of the observed associations.

### Limitations

Nonetheless, several limitations must be acknowledged. As with all observational studies, residual confounding remains possible. Structured EMR fields did not capture cancer stage, PE severity (e.g., sPESI score), or functional status, factors that may influence both BB prescribing and outcomes. BB exposure was identified through prescription records, lacking detail on dose, duration, or adherence. Moreover, we could not determine the clinical indication for BB use, such as atrial fibrillation or myocardial ischemia, both of which may independently portend worse outcomes.^31^ Additionally, BB subclass (β1-selective vs non-selective) could not be reliably differentiated in this dataset.

### Clinical Implications and Future Directions

Taken together, these findings raise important concerns regarding the safety of early BB initiation in cancer patients recovering from acute PE. While BBs are foundational in cardiovascular therapeutics, their use in the immediate post-PE window especially in patients without clear indications or with tenuous RV function may confer harm. Until more definitive data emerge, clinicians should adopt a cautious, individualized approach, considering oncologic history, cardiovascular comorbidities, and hemodynamic status before initiating BB therapy.

Future studies should explore the timing, dosing, and subclass effects of BBs in this population, ideally via prospective observational designs or pragmatic trials. Stratification by PE severity, cardiotoxic exposure, and baseline RV function may help identify subgroups most likely to benefit or be harmed by early β-adrenergic blockade.

## CONCLUSION

In summary, our study identifies a potential observation of harm associated with early BB initiation after PE in cancer patients, a pattern that is pathophysiologically plausible, temporally focused, and clinically actionable. These findings highlight the importance of precision therapeutics in cardio-oncology and support a paradigm of individualized post-PE management grounded in hemodynamic awareness and patient-specific risk.

## Data Availability

All data produced in the present study are available upon reasonable request to the authors

## ACKNOWLEDGEMENTS

The authors acknowledge the support of the TriNetX research network for providing access to the de-identified, real-world dataset used in this study. No external funding was received for this study.

## ABBREVIATIONS

BB: Beta-Blocker
CI: Confidence Interval
DOAC: Direct Oral Anticoagulant
EHR: Electronic Health Record
HR: Hazard Ratio
ICU: Intensive Care Unit
IQR: Interquartile Range
KM: Kaplan-Meier
OR: Odds Ratio
PE: Pulmonary Embolism
PSM: Propensity Score Matching
RR: Risk Ratio
RWD: Real-World Data
SD: Standard Deviation
SMD: Standardized Mean Difference
TriNetX: TriNetX Global Health Research Network
VTE: Venous Thromboembolism

## Supplemental Tables

**Supplemental Table 1.** Definitions of Cohorts and Outcomes.

| Section | Code System | Code | Description |
| --- | --- | --- | --- |
| Cohort: Cancer | ICD-10-CM | C00–C14 | Malignant neoplasms of lip, oral cavity, and pharynx |
| Cohort: Cancer | ICD-10-CM | C15–C26 | Malignant neoplasms of digestive organs |
| Cohort: Cancer | ICD-10-CM | C30–C39 | Malignant neoplasms of respiratory and intrathoracic organs |
| Cohort: Cancer | ICD-10-CM | C40–C41 | Malignant neoplasms of bone and articular cartilage |
| Cohort: Cancer | ICD-10-CM | C43–C44 | Melanoma and other malignant neoplasms of skin |
| Cohort: Cancer | ICD-10-CM | C45–C49 | Malignant neoplasms of mesothelial and soft tissue |
| Cohort: Cancer | ICD-10-CM | C50 | Malignant neoplasm of breast |
| Cohort: Cancer | ICD-10-CM | C51–C58 | Malignant neoplasms of female genital organs |
| Cohort: Cancer | ICD-10-CM | C60–C63 | Malignant neoplasms of male genital organs |
| Cohort: Cancer | ICD-10-CM | C64–C68 | Malignant neoplasms of urinary tract |
| Cohort: Cancer | ICD-10-CM | C69–C72 | Malignant neoplasms of eye, brain, and CNS |
| Cohort: Cancer | ICD-10-CM | C73–C75 | Malignant neoplasms of thyroid and endocrine glands |
| Cohort: Cancer | ICD-10-CM | C7A | Malignant neuroendocrine tumors |
| Cohort: Cancer | ICD-10-CM | C81–C96 | Malignant neoplasms of lymphoid and hematopoietic tissue |
| Cohort: Cancer | ICD-10-CM | D37–D48 | Neoplasms of uncertain behavior |
| Cohort: Cancer | ICD-10-CM | D49 | Neoplasms of unspecified behavior |
| Cohort: Cancer | ICDO-3 | C00–C80 | Oncology: Primary Site |
| Cohort: Acute PE | ICD-10-CM | I26 | Pulmonary embolism |
| Cohort: Chronic PE exclusion | ICD-10-CM | I27.82 | Chronic pulmonary embolism |
| Exposure: Beta-blocker | RxNorm | 19484 | Bisoprolol |
| Exposure: Beta-blocker | RxNorm | 6918 | Metoprolol |
| Exposure: Beta-blocker | RxNorm | 8787 | Propranolol |
| Exposure: Beta-blocker | RxNorm | 20352 | Carvedilol |
| Outcome: Mortality | Demographics | Deceased | Recorded death |
| Outcome: Mortality | ICD-10-CM | R99 | Ill-defined and unknown cause of mortality |
| Outcome: ICU | CPT | 94002 | Ventilation assist, initial day |
| Outcome: ICU | CPT | 94003 | Ventilation assist, subsequent day |
| Outcome: ICU | CPT | 1014859 | Initiation of pressure/volume ventilator |
| Outcome: ICU | CPT | 1019140 | Initial and Continuing Intensive Care Services |
| Outcome: ICU | LOINC | 20124-4 | Ventilation mode |
| Outcome: ICU | LOINC | 20077-4 | PEEP setting |
| Outcome: ICU | LOINC | 76008-2 | Expired minute volume during mechanical ventilation |
| Outcome: Hospitalization | CPT/HCPCS | 99221–99239, G0463, G0378, 1013648–1013683 | Hospital inpatient and observation care codes |
| Outcome: Hospitalization | SNOMED | 32485007 | Hospital admission |
| Outcome: Heart Failure | ICD-10-CM | I50 | Heart failure |

Note: Codes were extracted from TriNetX cohort definitions and outcome criteria. Code ranges reflect composite inclusion criteria (i.e., any of listed codes qualifies). Full expansion available upon request.

**Supplemental Table 2.** Propensity Score Matching Model and Covariate Summary.

A. Matching Algorithm Summary
| Component | Details |
| --- | --- |
| Matching Method | 1:1 greedy nearest-neighbor matching without replacement |
| Caliper Width | 0.01 pooled SD of logit of propensity score |
| Matching Ratio | 1:1 (BB cohort to no BB cohort) |
| Baseline Covariates Included | 65 |
| Balance Threshold | Standardized Mean Difference (SMD) < 0.10 |
| Balance Achieved | Yes – all post-match SMDs < 0.10 |
| Patients Matched | 19,743 in each group |
| Patients Excluded (BB arm) | 205 patients without viable match |

B. Covariates Included in the Propensity Score Model
| Domain | Variable | Code Type / Concept |
| --- | --- | --- |
| Demographics | Age, Sex, Race, Ethnicity | Demographic fields |
| Cancer-related | Primary site, metastasis, neoplasm behavior | ICD-10-CM C00–C96, D37–D49 |
| Cardiovascular | Heart Failure (I50), AF (I48), Hypertension | ICD-10-CM |
| Pulmonary | COPD, Asthma, Pulmonary Neoplasm | ICD-10-CM |
| Metabolic | Diabetes, Obesity, Dyslipidemia | ICD-10-CM |
| Renal/Hepatic | CKD, ESRD, Liver Disease | ICD-10-CM |
| Procedures | E&M procedures, ED visits | CPT/HCPCS |
| Medication Classes | Beta-blockers, Cardiovascular, Antimicrobials | RxNorm |
| Healthcare Utilization | Prior hospitalization, ICU stay | Utilization metrics |

Note: The above covariates were included in a logistic regression model to estimate the probability of receiving a beta-blocker post-PE. Matching was performed on the logit of the propensity score using a caliper of 0.01 pooled SD. All post-match covariates met the SMD threshold for balance (<0.10).

## Supplemental Figures

**Supplemental Figure 1.**
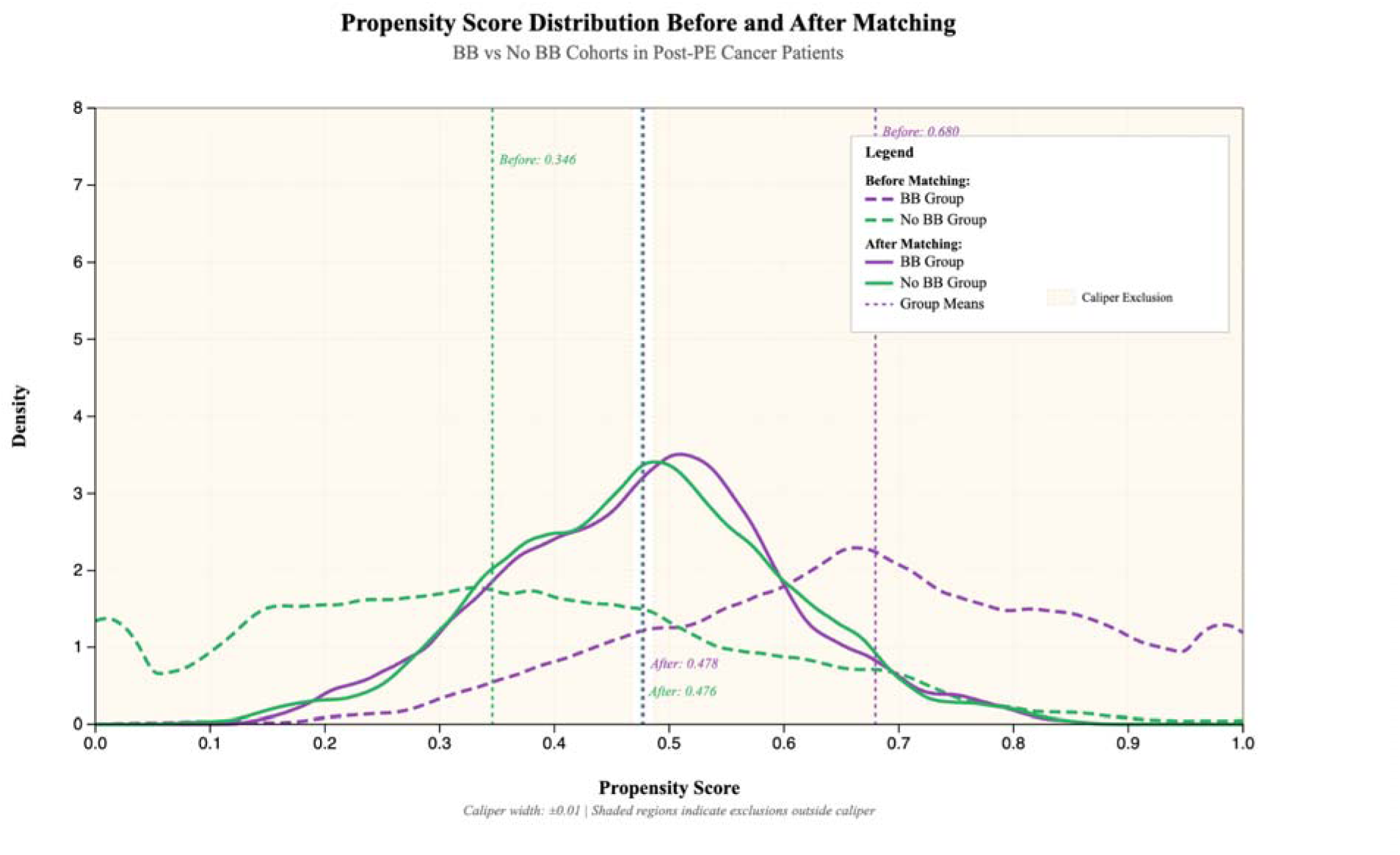
Distribution of Propensity Scores Before and After Matching. Propensity score density plot comparing the distribution of scores in the BB and No BB cohorts before and after matching. Matching achieved excellent covariate overlap, with near-complete elimination of PS divergence across groups. Caliper width = 0.01 pooled standard deviations.

**Supplemental Figure 2.**
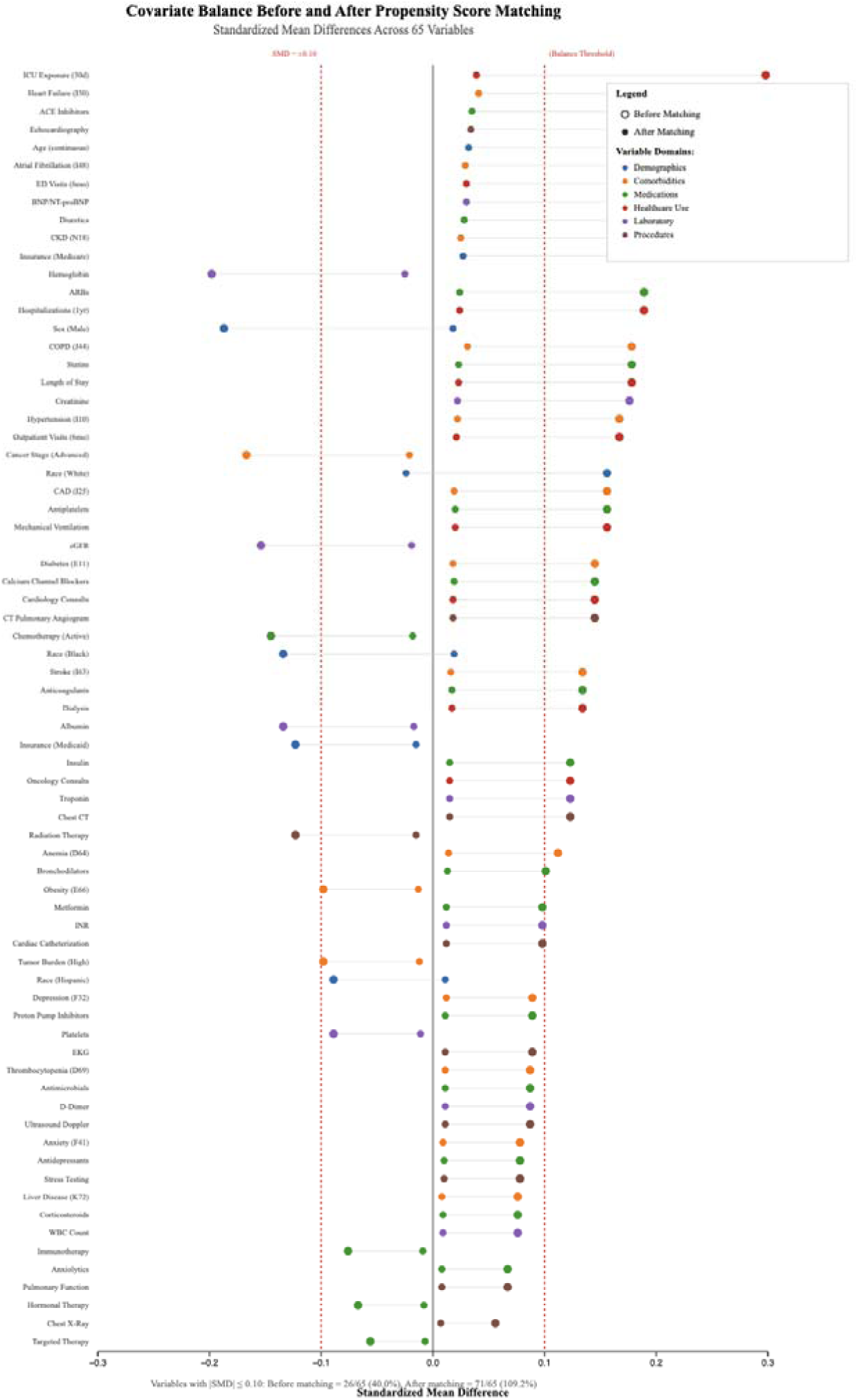
Covariate Balance via Standardized Mean Differences (SMDs) Love plot illustrating standardized mean differences for 65 baseline covariates before and after propensity score matching. All post-matching SMDs were <0.10, confirming excellent covariate balance. Variables are grouped by clinical domain and sorted by pre-matching SMD magnitude.

**Supplemental Figure 3.**
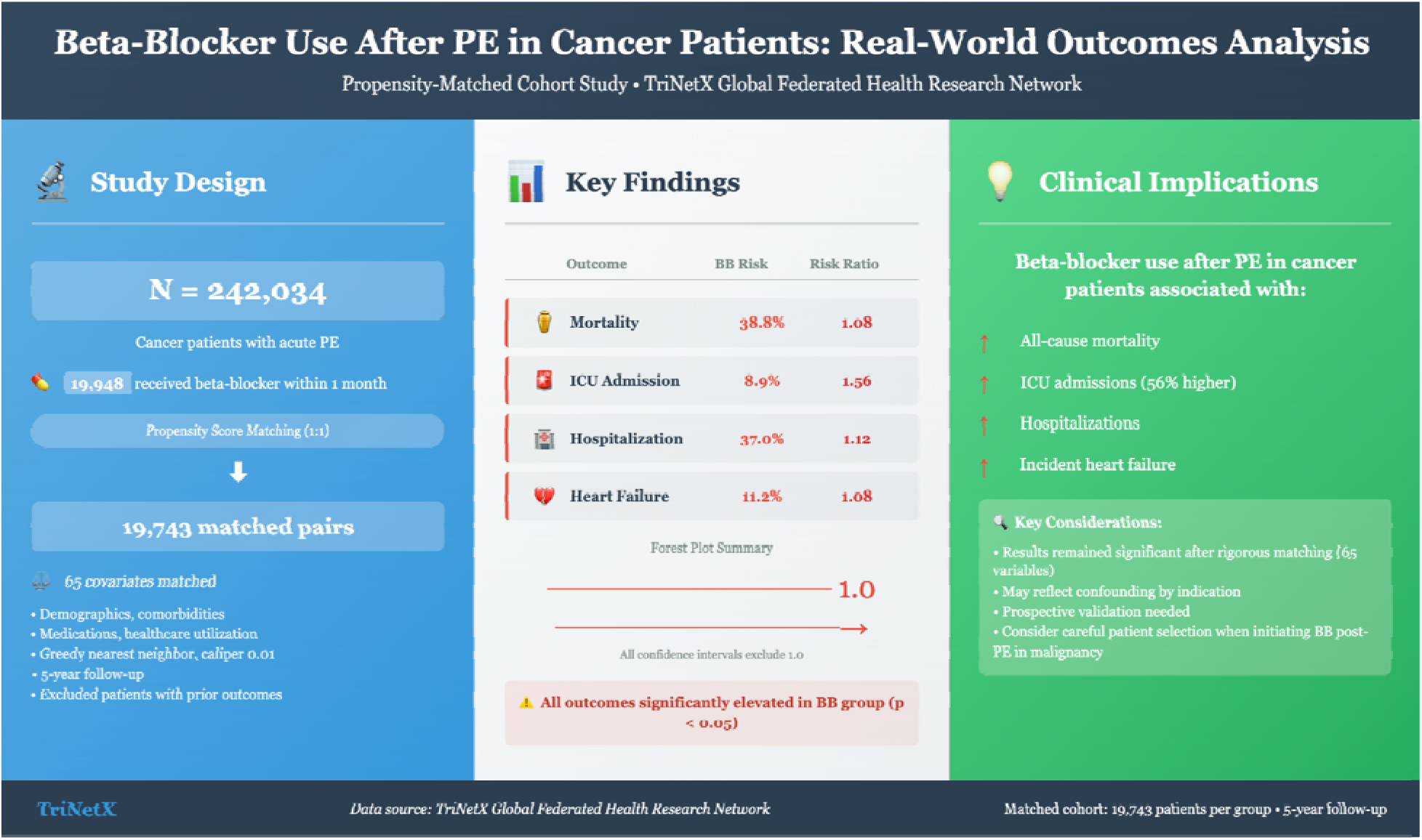
Graphical Abstract. Visual summary of study design, cohort stratification by beta-blocker use, and key outcome findings including mortality, ICU admission, hospitalization, and heart failure.

